# Changes in Transmission and Symptoms of SARS-CoV-2 in United States Households, April 2020–September 2022

**DOI:** 10.1101/2023.05.18.23290185

**Authors:** Alexandra M. Mellis, Adam S. Lauring, H. Keipp Talbot, Huong Q. McLean, Kerry Grace Morrissey, Melissa S. Stockwell, Natalie M. Bowman, Yvonne Maldonado, Katherine D. Ellingson, Suchitra Rao, Jessica E. Biddle, Sheroi Johnson, Constance Ogokeh, Phillip P. Salvatore, Carrie Reed, Sarah E. Smith-Jeffcoat, Jennifer K. Meece, Kayla E. Hanson, Edward A. Belongia, Emily E. Bendall, Julie Gilbert, Vanessa Olivo, Lori S. Merrill, Son H. McLaren, Ellen Sano, Celibell Y. Vargas, Lisa Saiman, Raul A Silverio Francisco, Ayla Bullock, Jessica Lin, Prasanthi Govindarajan, Sarah H. Goodman, Clea C. Sarnquist, Karen Lutrick, Karla I. Ledezma, Ferris A. Ramadan, Kathleen Pryor, Flavia N Miiro, Edwin Asturias, Samuel Dominguez, Daniel Olson, Hector S. Izurieta, James Chappell, Christopher Lindsell, Natasha Halasa, Kimberly Hart, Yuwei Zhu, Jonathan Schmitz, Melissa A. Rolfes, Carlos G. Grijalva, the RVTN Study Group

## Abstract

**Background:** The natural history of SARS-CoV-2 infection and transmission dynamics may have changed as SARS-CoV-2 has evolved and population immunity has shifted.

**Methods:** Household contacts, enrolled from two multi-site case-ascertained household transmission studies (April 2020–April 2021 and September 2021–September 2022), were followed for 10–14 days after enrollment with daily collection of nasal swabs and/or saliva for SARS-CoV-2 testing and symptom diaries. SARS-CoV-2 virus lineage was determined by whole genome sequencing, with multiple imputation where sequences could not be recovered. Adjusted infection risks were estimated using modified Poisson regression.

**Findings:** 858 primary cases with 1473 household contacts were examined. Among unvaccinated household contacts, the infection risk adjusted for presence of prior infection and age was 58% (95% confidence interval [CI]: 49–68%) in households currently exposed to pre-Delta lineages and 90% (95% CI: 74–100%) among those exposed to Omicron BA.5 (detected May – September 2022). The fraction of infected household contacts reporting any symptom was similarly high between pre-Delta (86%, 95% CI: 81–91%) and Omicron lineages (77%, 70–85%). Among Omicron BA.5-infected contacts, 48% (41–56%) reported fever, 63% (56–71%) cough, 22% (17–28%) shortness of breath, and 20% (15–27%) loss of/change in taste/smell.

**Interpretation:** The risk of infection among household contacts exposed to SARS-CoV-2 is high and increasing with more recent SARS-CoV-2 lineages. This high infection risk highlights the importance of vaccination to prevent severe disease.

**Funding:** Funded by the Centers for Disease Control and Prevention and the Food and Drug Administration.

**Key points:**

- Monitoring the transmissibility and symptomatology of SARS-CoV-2 lineages is important for informing public health practice and understanding the epidemiology of COVID-19; household transmission studies contribute to our understanding of the natural history of SARS-CoV-2 infections and the transmissibility of SARS-CoV-2 variants.
- The Omicron BA.5 sub-lineage is highly transmissible, similar to previous Omicron sub-lineages.
- Over 80% of infected household contacts reported at least 1 symptom during their infection and the proportion of household contacts with asymptomatic infection did not differ by SARS-CoV-2 variant. The most common symptom was cough. Change in taste or smell was more common in Omicron BA.5 infections, compared to previous Omicron sub-lineages, but less common compared to pre-Delta lineages.
- The high infection risk among household contacts supports the recommendations that individuals maintain up-to-date and lineage-specific vaccinations to mitigate further risks of severe disease.

## Background

As novel SARS-CoV-2 lineages have emerged, changes in transmissibility have been monitored through population-level surveillance combined with understanding of the biological features of the new viruses^1^. Omicron BA.5 was first observed in the United States in May of 2022, showing a substantial transmission advantage over other lineages circulating at the time^2,3^ and improved ability to escape immunity from SARS-CoV-2 vaccines^4,5^ and from immunity induced from previous infection with circulating SARS-CoV-2 viruses, including prior Omicron lineages^6^. These viral variants also emerged in the context of greater population immunity due to vaccination and prior infections, although substantial heterogeneity exists in individual-level protection. Changes in COVID-19 severity with emerging virus variants have also been monitored primarily through cross-sectional surveillance^7^. However, these approaches have not characterized important changes in symptomatic proportions and transmissibility over time.

The Respiratory Virus Transmission Network is an ongoing household transmission study recruiting index cases and their household contacts at sentinel sites nationwide in the United States. The prospective close follow-up of households with a known exposure to SARS-CoV-2 allows for examination of infection risk among household contacts, characterization of the history of natural infection and vaccination, insight into the timing of transmission events, and description of the behavioral changes in infection mitigation measures within household settings. Here, we combine data from the Respiratory Virus Transmission Network and a previous SARS-CoV-2 household transmission study to compare these features across multiple periods of SARS-CoV-2 circulation from pre-delta through Omicron BA.5 lineage viruses.

## Methods

### Study population

Case-ascertained household transmission studies with harmonized protocols were conducted at multiple sites across the United States April 2020 – April 2021 (hereafter, the “early pandemic study”; see^8-10^) and September 2021 – September 2022 (hereafter, the “ongoing study”; **Figure 1**). Individuals who tested positive for SARS-CoV-2 on either molecular assay or rapid antigen testing were identified as index cases by clinical testing at academic medical centers, health systems, or public health registries (static testing sites; in the early pandemic study and the ongoing study), or via online recruitment through remote testing services, participatory surveillance systems, or other commercial testing platforms (in the ongoing study only). Households were screened and enrolled within 6 days of the earliest symptom onset in the household, and informed consent was obtained from index cases and their household contacts. At each participating research site, IRBs approved study protocols. CDC determined these activities were conducted consistent with applicable federal law and CDC policy (see 45 C.F.R. part 46; 21 C.F.R. part 56).

**Figure 1.**
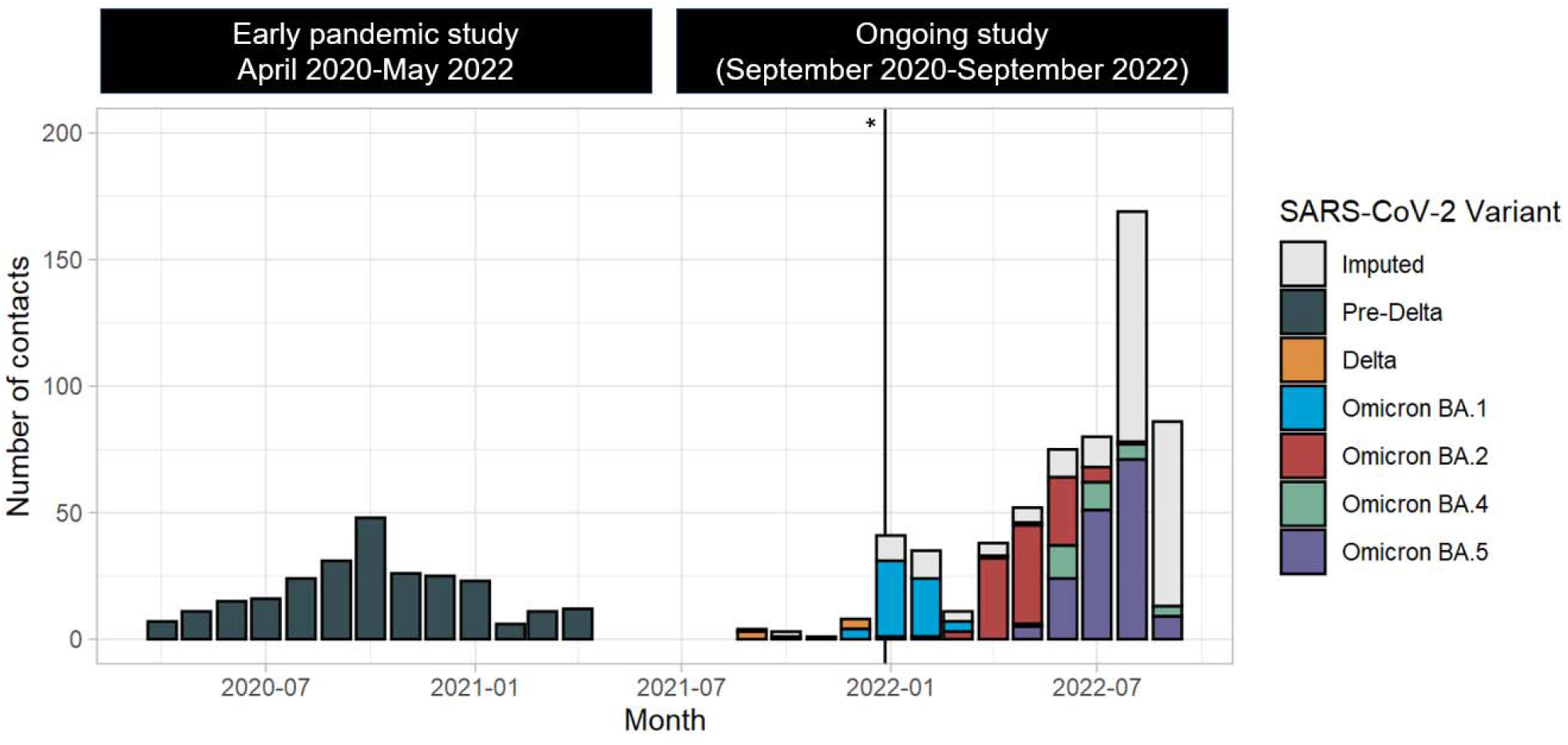
SARS-CoV-2 household transmission study enrollment in the United States by date of first illness onset in the household and viral variant, April 2020-September 2022. Bars show the number of household contacts enrolled by month of household exposure, defined as the date of first symptom onset or positive test in the household, by the SARS-CoV-2 variant circulating in the household. If all sequencing information was missing, the variant was imputed in subsequent analyses. The asterisk and vertical bar indicate the timing of public health guidance change for the isolation of cases on December 27, 2021.

At enrollment, participants self-reported demographics, household characteristics, vaccination status, and previous SARS-CoV-2 infection history. All participants were prospectively followed for 14 days (in the early pandemic study) or 10 days (in the ongoing study), including symptom diaries (assessing aches, fatigue, fever, vomiting, abdominal pain, diarrhea, cough, chest tightness/pain, shortness of breath, wheezing, headache, change of taste/smell, sore throat, nasal congestion, and runny nose), daily logs of infection control practices, and daily self- or parent-collection of nasal or saliva samples. In the ongoing study, nasal samples were collected among households recruited from static testing sites, and saliva samples were collected among households recruited online. In the early pandemic study, nasal or saliva samples were collected among households recruited at Vanderbilt University Medical Center and nasal samples were collected among households recruited at Marshfield Clinic Health System.

Five infection control practices were assessed in daily logs, including whether the participant slept in a separate bedroom from others; used a separate bathroom; ate separately; spent under an hour in the same room as others; and used a mask when sharing a room. Participants were considered to be fully isolated if they used all practices for five days after their symptom onset Throughout 2020 and until December 27, 2021, CDC recommended full isolation from others for ten days following symptom onset; this was subsequently shortened to five days, with mask-wearing recommended for an additional five days^11^. In the early pandemic study, infection control practices were assessed only among index cases (regardless of primary case status) recruited November 2020. In the ongoing study, infection control practices were assessed among all participants.

Study staff verified vaccination status using vaccination cards, state vaccination registries, and medical records. Vaccination status was determined based on the number of doses reported by either plausible self-report (report of a manufacturer plus either the date or location of vaccine receipt) or by documentation through the vaccine verification process. Only doses received more than 14 days prior to the earliest symptom onset in the household were considered for vaccination status determination. Individuals who received only a single dose of an mRNA vaccine were combined with those who had received no doses in analyses.

### Laboratory methods

Daily respiratory samples underwent testing by molecular assay for qualitative detection of SARS-CoV-2. In the ongoing study, saliva samples were tested by real-time reverse-transcription polymerase chain reaction (RT-PCR) using the Sampled testing platform (EUA: 137776), and nasal samples were tested by transcription mediated amplification (TMA) using the Panther Fusion Hologic system. In the ongoing study, one positive specimen per person underwent whole-genome sequencing. Sequencing was performed by the University of Michigan (which received the first specimen with an RT-PCR cycle (CT) threshold value under 32 for static testing site recruitment) or Sampled/Infinity Biologix (which received the first specimen with an RT-PCR CT value under 30 for remote recruitment). Lineages were assigned using PANGO (version 3.1.20 – 4.1.2)^12^ and Nextclade^13^, and mapped to variants (Delta, Omicron BA.1, Omicron BA.2, Omicron BA.4, or Omicron BA.5; see **Supplementary Table 1**). In the early pandemic study, nasal or saliva samples were tested by RT-PCR using ThermoFisher, Quidel, or CDC-developed assays, without sequencing.

### Statistical analysis

We considered the primary case in each household to be the individual who tested positive for SARS-CoV-2 with the earliest evidence for infection, either the first symptom onset or the first positive test if all cases were asymptomatic. Households were excluded for having multiple primary cases if multiple individuals were infected and had symptom onset on the same date (**Figure 2**). Household contacts were considered eligible for analysis if they collected respiratory samples on at least 7 days, and their vaccination status was known. Only contacts who were age-eligible to receive a booster vaccination dose (over the age of 5 years) were included. Individuals who received only a single dose of Johnson & Johnson, the viral vector vaccine, were excluded due to sample size (**Figure 2**). Levels of vaccination for this analysis were that the contact had received 0-1 dose of mRNA vaccine, or, relative to earliest onset in the household received 2 doses of vaccine 120 days prior, received 2 doses less than 120 days prior, received 3 doses 120 days or more prior, received 3 doses less than 120 days prior, or received 4 doses.

**Figure 2.**
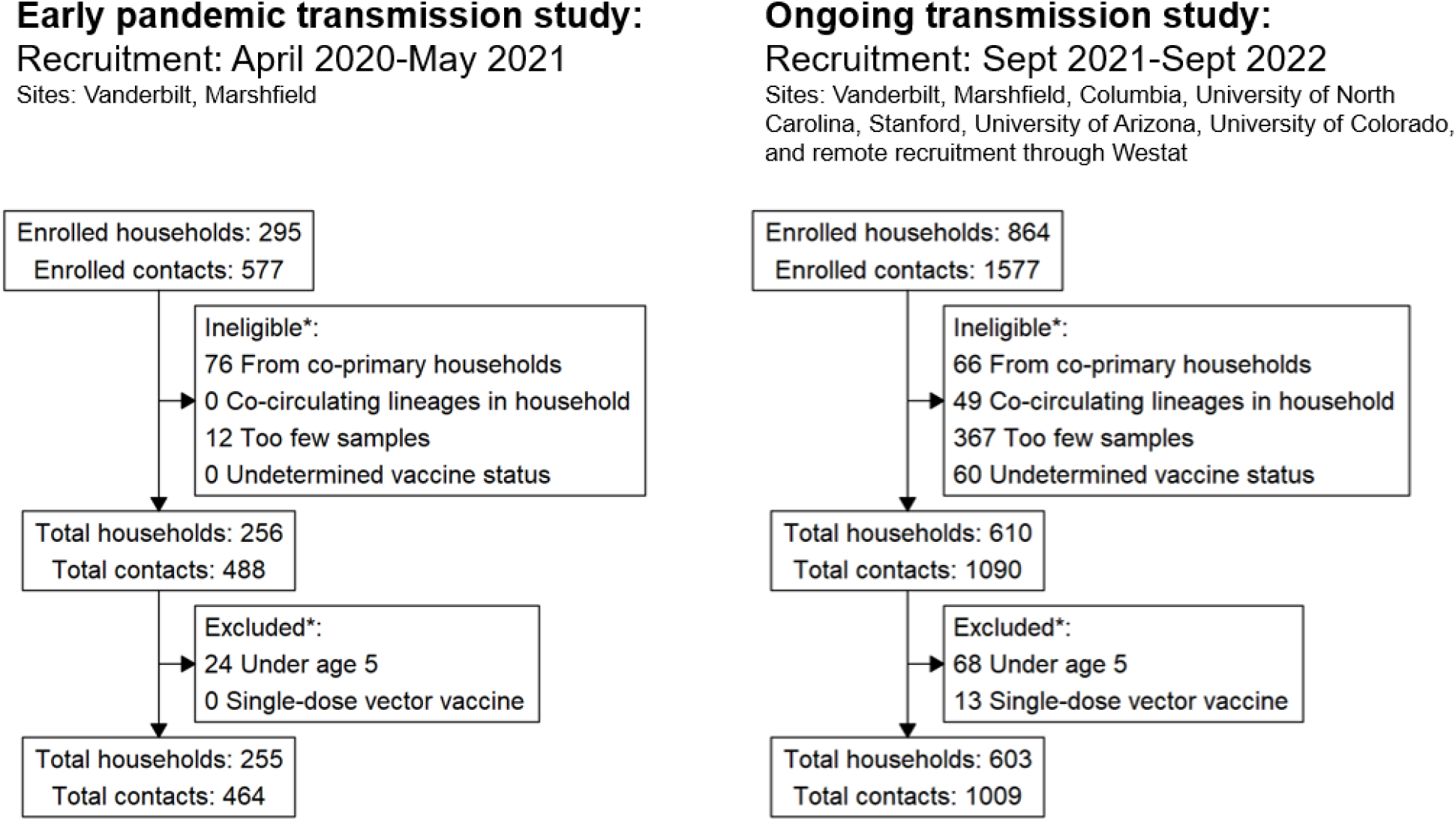
Selection criteria for two household transmission studies, United States, 2020-2022. Footnote: *Participants could be ineligible or excluded for multiple reasons.

SARS-CoV-2 lineage assignments were made at the household level. In the early pandemic transmission study, all households were assumed to be exposed to pre-Delta variants. In the ongoing investigation, households were classified by PANGO lineages from valid sequencing assignments in the household, and households with co-circulation of multiple lineages were excluded (**Figure 2**). Sequences from households recruited from static testing sites were also manually reviewed for minor changes by one author, AL, to indicate co-circulation^14^. When no valid sequencing results were available, lineage was multiply imputed based on the variant proportions in the Health and Human Services (HHS) region^15^ and at the time of first symptoms in the household.

Primary outcomes were the risk of infection (detection of any SARS-CoV-2 positivity) among contacts, the symptomatic fraction among infected cases, the serial interval (time from symptom onset in the primary case to symptom onset among infected contacts), and the infection control practices of primary cases in the households. SARS-CoV-2 detections were pooled across nasal and saliva specimens^16^. Infection risk was estimated using multivariable modified Poisson regression^17^ including SARS-CoV-2 variant, age (under 18, 18-64, and over 65 years of age) and self-reported prior infection, and accounting for household clustering using generalized estimating equations (GEE). Models were stratified by vaccination, to compare infection risks by lineage among similar vaccination histories. Symptomatic fractions were compared using logistic regression by SARS-CoV-2 lineage, using GEEs for household clustering. Serial intervals among symptomatic primary case-infected contact pairs were calculated for each SARS-CoV-2 lineage using the EpiEstim package^18^. Estimates for regressions and serial interval distributions were pooled across five multiply imputed datasets using Rubin’s rules. The infection control practices of primary cases were compared across guidance periods, for symptomatic primary cases with symptom onset on or after December 27, 2021, compared to those with symptom onset prior to this change in guidance using univariate logistic regression.

## Results

During the two study time periods, a total of 1109 households were enrolled. Of these, 858 single-primary case households were eligible for this analysis (255 households with 464 contacts enrolled from April 2020–April 2021 and 603 households with 1009 contacts enrolled from September 2021– September 2022). Eligible contacts ranged in age from 5 to 85 years (27% under 18 years, 64% between 18 and 64 years, and 9% 65 years of age and older; **Table 1**). All individuals in the early pandemic study had received 0 or 1 dose of vaccine. In the ongoing study, 137 (14%) of contacts had received 0 or 1 dose, 245 (24%) had received 2 doses 120 days or more prior to household exposure, 43 (4%) had received 2 doses within 120 days, 360 (36%) had received 3 doses 120 or more days prior, 113 (11%) had received 3 doses within 120 days, 111 (11%) had received 4 doses. Only 28 (3%) of the eligible contacts had received a mixed vaccine series including both adenoviral vector and mRNA products. In the ongoing study, 29% of contacts reported a prior SARS-CoV-2-positive test, a median of 213 days (interquartile range: 85–476) days before earliest symptom onset in their household.

**Table 1.**
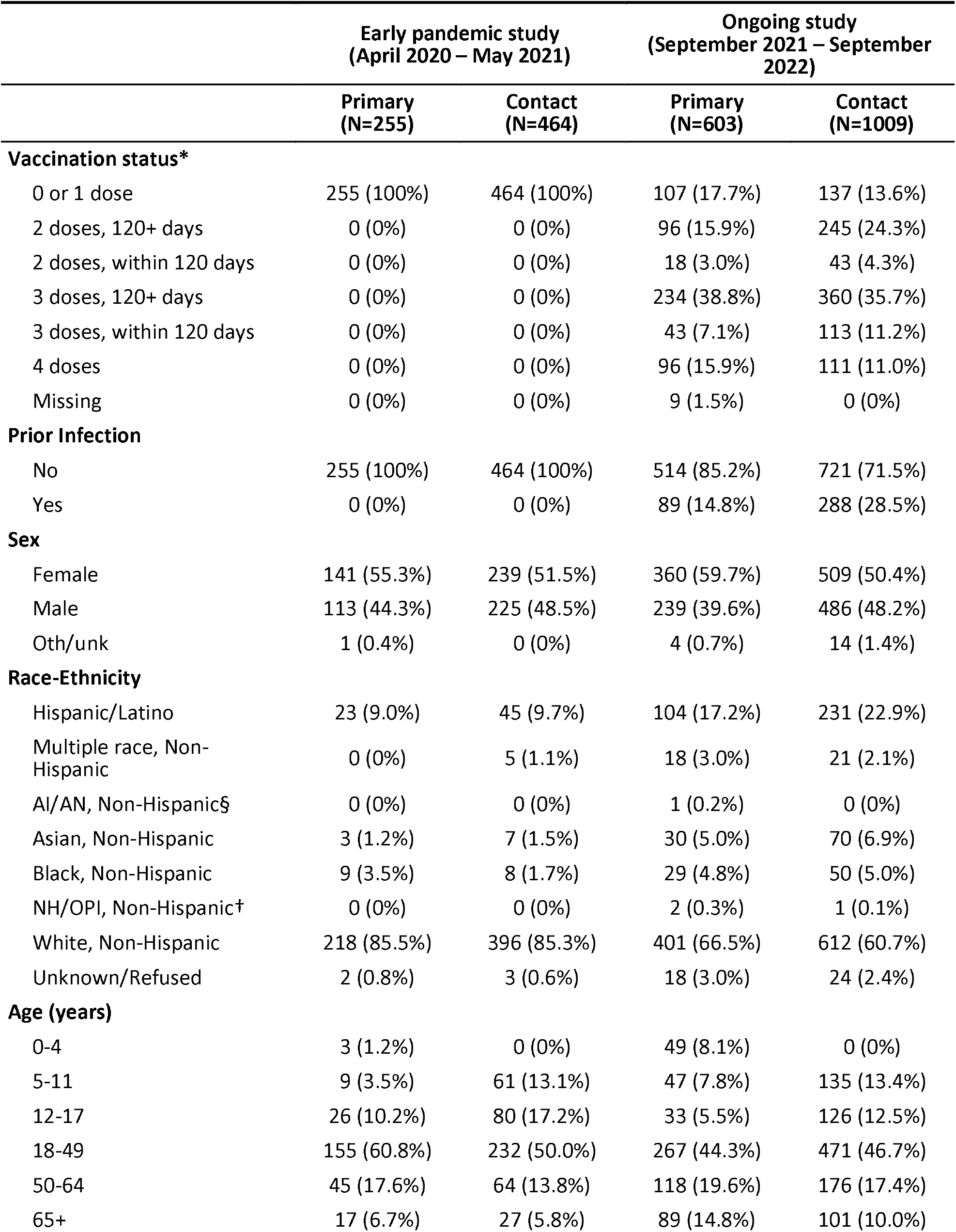
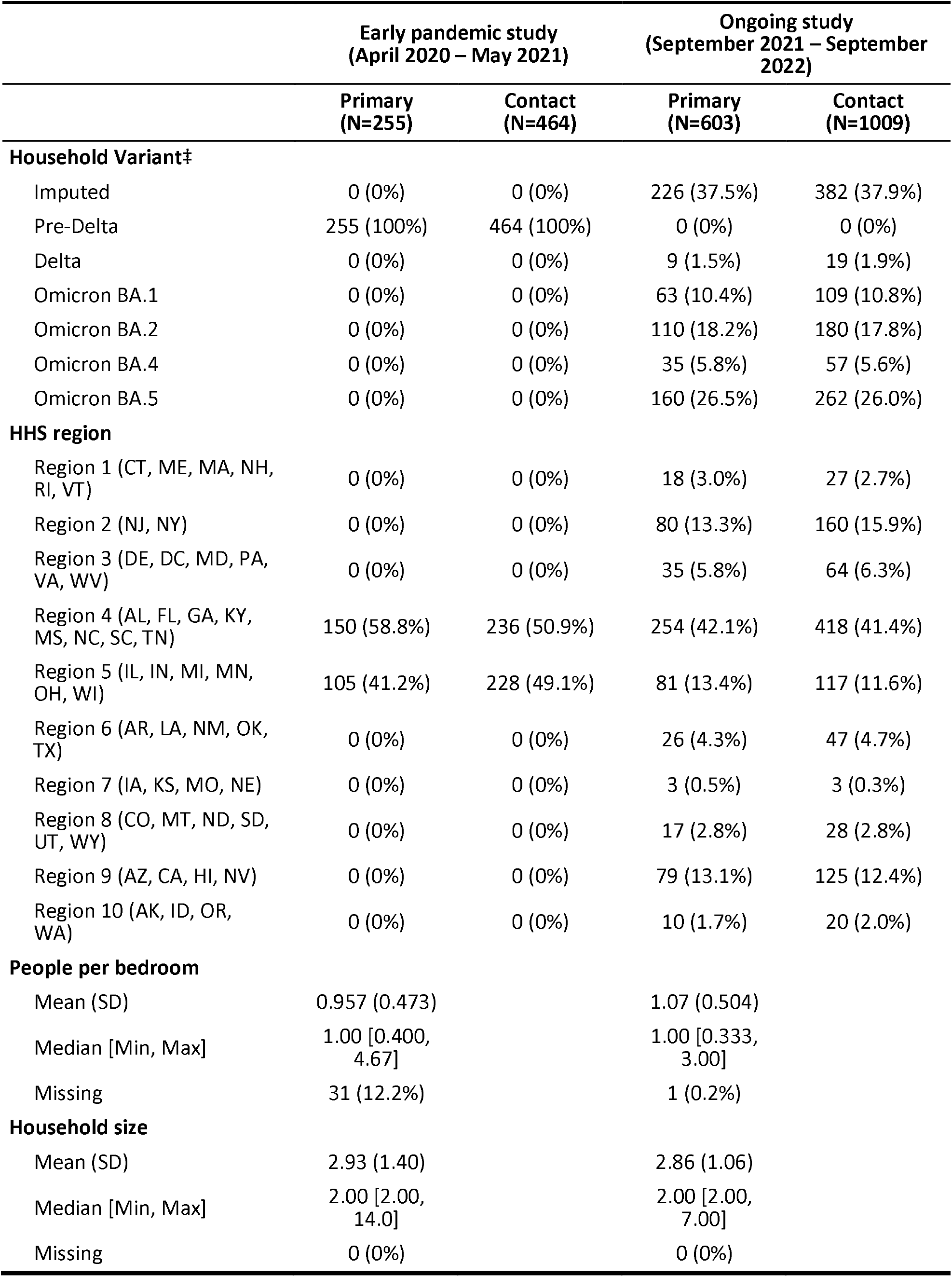
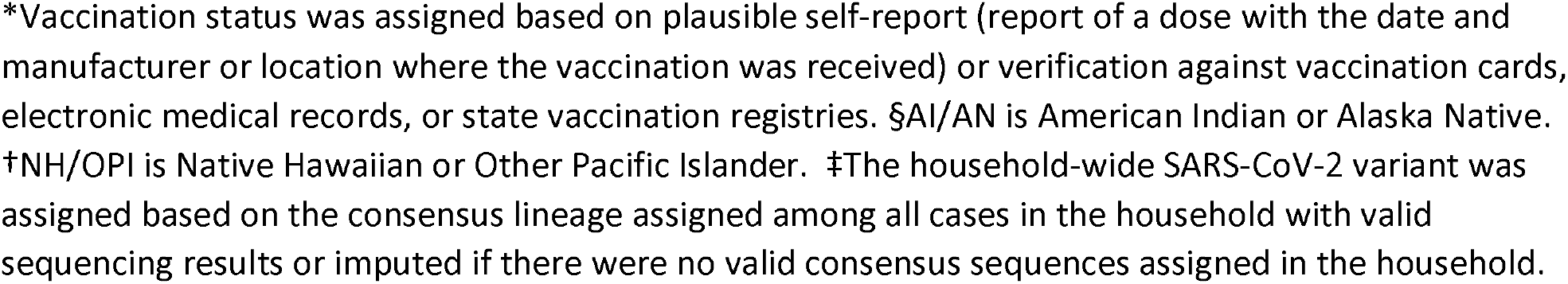
Characteristics of primary cases and household contacts from two case-ascertained SARS-CoV-2 household transmission studies, United States, 2020-2022.

Valid SARS-CoV-2 sequence assignments were available for most households in the ongoing study, but variants were imputed for 37% of households. The crude risk of SARS-CoV-2 infection among contacts ranged from 47% to 76% across variants of the virus (**Table 2**). The serial interval ranged from 3.8 days (pooled 95% uncertainty interval [UI]: 3.4–4.1 days) among contacts exposed to Omicron BA.5 to 4.7 days (95%UI: 4.4–5.0 days) among pre-Delta household contacts; serial interval estimates were not generated for Delta-exposed contacts due to small sample size. The fraction of infected contacts reporting any symptom was similar across lineages, but fever was more frequently reported among Omicron BA.5-infected contacts (48%, 95% CI: 41–56%) compared to pre-Delta-infected contacts (37%, 95% CI: 31–44%), and change of taste or smell was less frequently reported (20% [95% CI: 15–27%] among Omicron BA.5-infected contacts, compared to 42% [95% CI: 36–49%] among pre-Delta-infected contacts; **Supplementary Figure 1**).

**Table 2.**
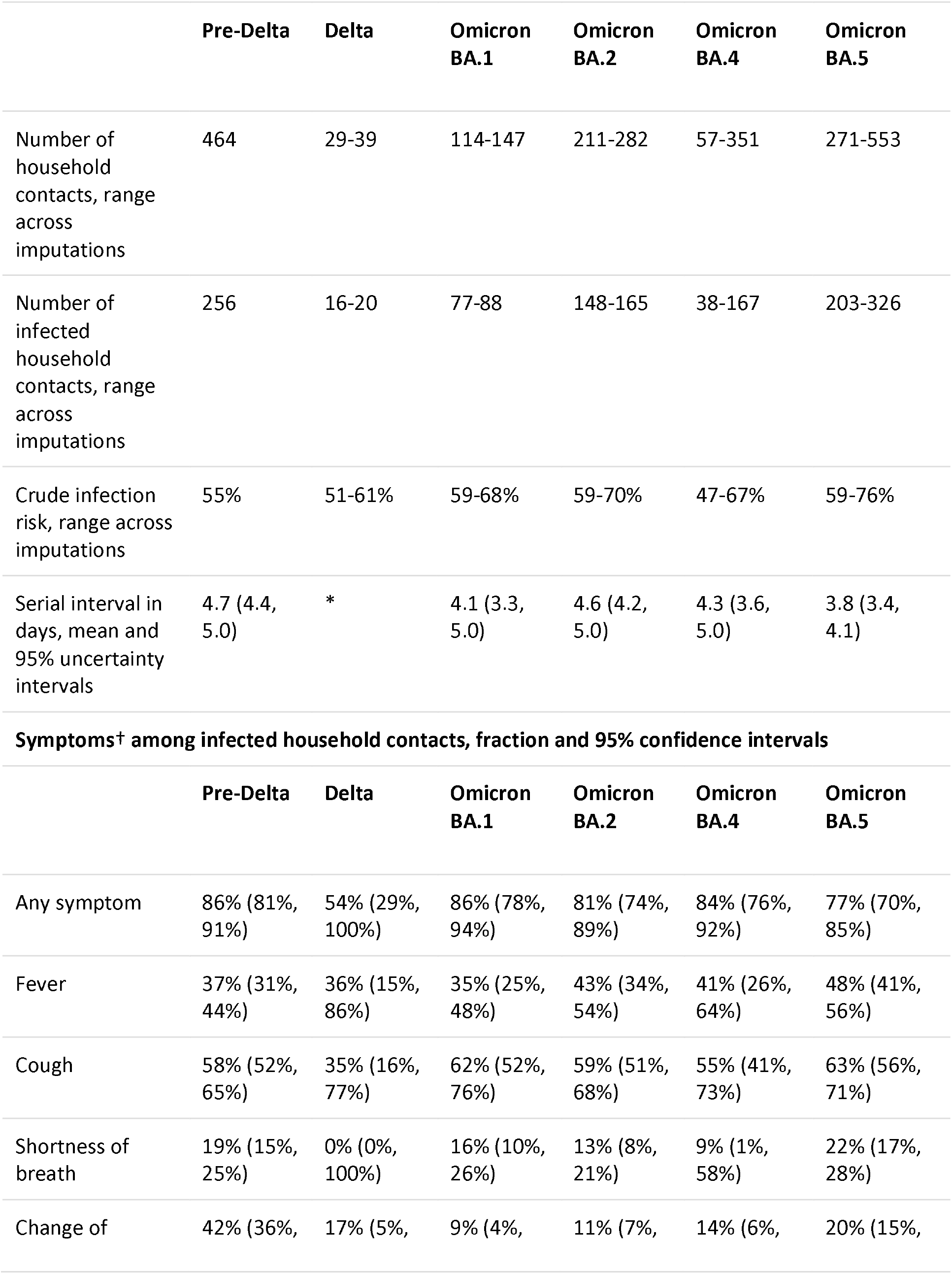
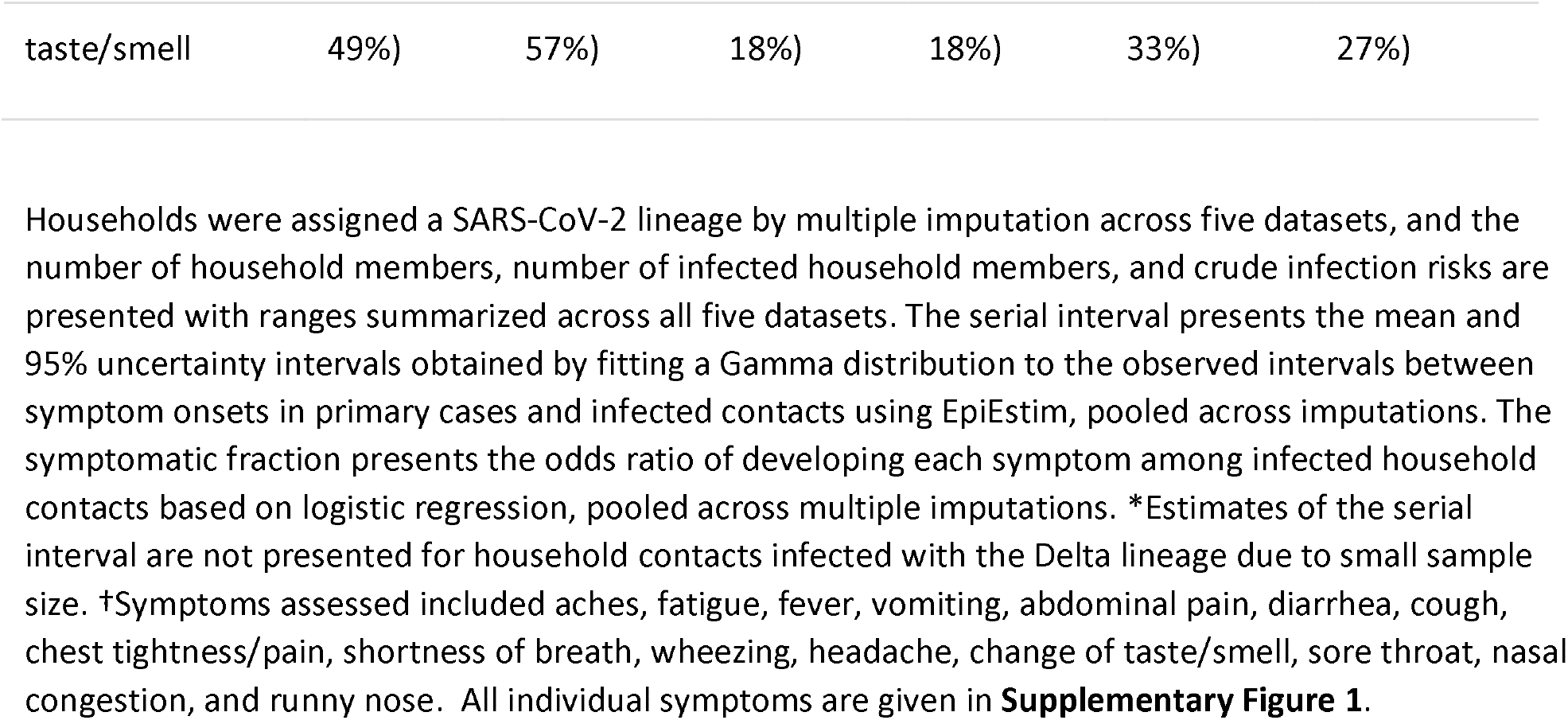
Infections and symptoms among household contacts by SARS-CoV-2 variant, United States, 2020-2022.

|  | Pre-Delta | Delta | Omicron<br>BA.1 | Omicron<br>BA.2 | Omicron<br>BA.4 | Omicron<br>BA.5 |
| --- | --- | --- | --- | --- | --- | --- |
| Number of household contacts, range across imputations | 464 | 29-39 | 114-147 | 211-282 | 57-351 | 271-553 |
| Number of infected household contacts, range across imputations | 256 | 16-20 | 77-88 | 148-165 | 38-167 | 203-326 |
| Crude infection risk, range across imputations | 55% | 51-61% | 59-68% | 59-70% | 47-67% | 59-76% |
| Serial interval in days, mean and 95% uncertainty intervals | 4.7 (4.4, 5.0) | * | 4.1 (3.3, 5.0) | 4.6 (4.2, 5.0) | 4.3 (3.6, 5.0) | 3.8 (3.4, 4.1) |
| **Symptoms<sup>†</sup> among infected household contacts, fraction and 95% confidence intervals** |  |  |  |  |  |  |
|  | Pre-Delta | Delta | Omicron<br>BA.1 | Omicron<br>BA.2 | Omicron<br>BA.4 | Omicron<br>BA.5 |
| Any symptom | 86% (81%, 91%) | 54% (29%, 100%) | 86% (78%, 94%) | 81% (74%, 89%) | 84% (76%, 92%) | 77% (70%, 85%) |
| Fever | 37% (31%, 44%) | 36% (15%, 86%) | 35% (25%, 48%) | 43% (34%, 54%) | 41% (26%, 64%) | 48% (41%, 56%) |
| Cough | 58% (52%, 65%) | 35% (16%, 77%) | 62% (52%, 76%) | 59% (51%, 68%) | 55% (41%, 73%) | 63% (56%, 71%) |
| Shortness of breath | 19% (15%, 25%) | 0% (0%, 100%) | 16% (10%, 26%) | 13% (8%, 21%) | 9% (1%, 58%) | 22% (17%, 28%) |
| Change of | 42% (36%, | 17% (5%, | 9% (4%, | 11% (7%, | 14% (6%, | 20% (15%, |
| taste/smell | 49%) | 57%) | 18%) | 18%) | 33%) | 27%) |

The adjusted infection risk by SARS-CoV-2 lineage within each vaccination status stratum, accounting for age, prior SARS-CoV-2 infection, and household clustering, is presented in **Figure 3** and **Supplemental Table 2**. Among contacts who had received 0 or 1 dose of a vaccine, the overall risk of infection was 58% (95% CI: 49–68%) after pre-Delta exposure, 71% (95% CI: 33–100%) after Delta exposure, 79% (95% CI: 52%–100%) after Omicron BA.1 exposure, 77% (95% CI: 58%–100%) after Omicron BA.2 exposure, 68% (95% CI: 45%–100%) after Omicron BA.4 exposure, and 90% (95% CI: 74–100%) after Omicron BA.5 exposure. Among most contacts across vaccination strata and SARS-CoV-2 lineages, infection risks were above 50%. The precision of these estimates was limited, and confidence intervals of these infection risks overlapped among all groups (**Figure 3**).

**Figure 3.**
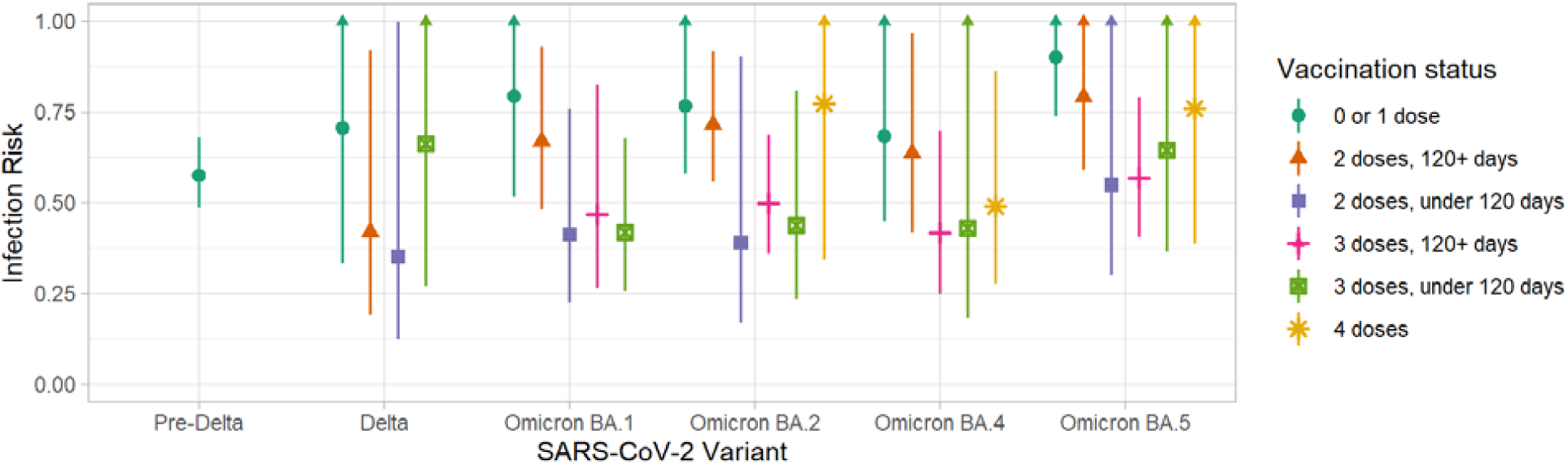
Infection risk among household contacts, by vaccination status and SARS-CoV-2 variant in the household, United States, 2020-2022. Point estimates show marginal infection risks for each SARS-CoV-2 lineage and vaccination status, and bars show 95% confidence intervals. Infection risks by variant were estimated by Poisson regressions stratified by the vaccination status of household contacts, accounting for age (under 18, 18-64, and over 65 years of age) and self-reported prior infection. Arrows indicate that the upper bound of these confidence intervals exceeded 1, a possible estimate in the modified Poisson regression chosen to model infection risk as a common outcome. Estimates were not generated for vaccination status levels that were only sparsely observed across imputed strata (e.g., individuals who were exposed to Omicron BA.1 were not included in the 4-dose vaccine model).

The infection control practices of primary cases were compared across time periods marked by changes in public health guidance for case isolation. Across both periods, it was infrequent for the primary case to isolate from household contacts for five days starting the day after symptom onset. In the earlier guidance period, 17 of 85 (20%) primary cases fully isolated for five days, compared to 62 of 550 (11%) primary cases in the later guidance period (rate ratio, 95% CI: 0.56, 0.35 – 0.95). Reports of individual practices (sleeping in a separate bedroom, eating separately, using a separate bathroom, wearing a mask if with others, and spending under an hour in the same room as others) were also marginally lower during the later guidance period. (**Supplementary Figure 2**).

## Discussion

Using data collected by household transmission studies conducted during the first two years of the COVID-19 pandemic, we explored the risk of SARS-CoV-2 transmission as novel viral variants emerged and as population immunity shifted. We observed consistently high infection risks to household contacts exposed to SARS-CoV-2 variants, declining serial intervals across emerging SARS-CoV-2 variants, high symptomatic fraction among infected contacts, and declining rates of infection control practices over successive variant waves. These case-ascertained household studies provide ideal platforms for monitoring the transmissibility of respiratory pathogens^19^, such as SARS-CoV-2. The prospective monitoring of household contacts also provides rich data for examining the natural history of SARS-CoV-2 infection, including changes in symptomatology as a function of both virus and host-level characteristics. These platforms have also allowed us to observe changes in reported use of mitigation behaviors in high-contact household settings, helping us better understand observed transmission dynamics within US households.

The risk of SARS-CoV-2 infection among household contacts was high with SARS-CoV-2 viruses circulating in 2020, with over 50% of contacts becoming infected, and has remained high in households impacted by Omicron lineage viruses, which circulated in late 2021 and 2022. This is consistent with population-level evidence of high Omicron infection rates, even with widespread vaccination, and with other reports of higher household attack rate or infection risk after Omicron exposures^20-22^. Other household studies have found high infection risks among contacts exposed to Omicron sub-lineages, at 81% of total and 86% of unvaccinated household contacts in a study with systematic contact testing^23^, and 29% of total and 28% of unvaccinated contacts in a study using household register data without systematic testing^24^. Estimates from registers with contact tracing but without systematic testing are intermediate, at approximately 51% of total contacts^25^. We observed a high burden of infection in contacts even among those with recent SARS-CoV-2 vaccination. This analysis, however, was not designed to assess vaccine effectiveness against infection and we cannot rule out other causes for our observations. We are also not able to make comparisons in rates of vaccination by household member vaccination status due to limited precision in these estimates.

The serial interval was similar among Omicron SARS-CoV-2 variant categories in our study. Among Omicron BA.5 infections, this interval was a day shorter than among pre-Delta infections. The estimated serial interval among these transmission pairs was slightly longer than that estimated in Song et al., at 2.9 days^20^, but consistent with other Omicron BA.1 estimates ^26-28^. Shorter serial intervals suggest changes in the incubation period or infectiousness profile, changes in interactions among household contacts, or a combination of these factors^29^. Our analyses did not interrogate the precise reasons of this observed shortening, but these data show that more recent Omicron sub-lineages may transmit with shorter serial intervals than pre-Delta lineages.

Population-level data have suggested that Omicron variant virus infections may be less severe than other SARS-CoV-2 infections^7,30^. In this study, the overall fraction experiencing any symptom was similar across different variants. The loss or change of taste and smell might be less frequent among Omicron lineage infections than among pre-Delta infections. While early reports found that loss of smell was less common during early Omicron periods than during Delta periods^31^, Google Search trends for taste loss have been more associated with Omicron BA.5 than prior Omicron lineages^3^ and an analysis of public health registry data^32^ reported that 19% of Omicron BA.2 infections and 25% of BA.5 infections experienced loss of taste.

In addition to changes in population immunity and virological differences, behavioral changes can influence differences in transmission. Public health guidance on the isolation of cases was shortened during the period of these studies. We observed that individuals who were the primary case in the household did not commonly isolate from other household members for five days in either guidance period. Reported use of these practices was also lower in households enrolled during the more recent guidance period. These practices may influence serial interval estimates and the frequent interactions in households may be contributing to the high infection risks observed across these studies.

There are several limitations to this analysis. First, the primary focus of this analysis was the impact of SARS-CoV-2 variant on infection risk, but sequencing data was not complete for all households enrolled in these studies. Rather than excluding these households (which may have biased estimates of the infection risk downwards, because households with fewer positive specimens might have been less likely to contribute sequences), data was multiply imputed using regional surveillance data. Second, the analysis of infection risk focused on the characterization of infections with different SARS-CoV-2 lineages stratified within vaccination status levels of household contacts but was not designed to generate direct estimates of the impact of vaccination on infection. We did not examine infection risks by several other key factors, such as individual histories of both infection and vaccination status or pre-existing conditions. Third, these household transmission studies primarily offer insights into mild illness in the community settings, but severe disease that required hospitalization was not assessed. Additionally, infection control practices were assessed by self-report, which may have introduced reporting bias. Finally, to examine a broader range of variants, data were pooled across studies with similar but not identical protocols; differences in study procedures like follow-up time, time to enrollment, and sample collection methods were not accounted for in final analyses due to structural confounding with observed variants.

Comparing the transmissibility and symptomatology of SARS-CoV-2 lineages across a common household transmission platform provides insights into the possibly changing epidemiology of COVID-19. This study highlights the need for continued surveillance to identify and monitor novel variants, and for measures such as receipt of recommended vaccinations to protect against severe disease given high infection risks.

## Data Availability

All data produced in the present study are available upon reasonable request to the authors

## Acknowledgements

The authors would like to acknowledge all members of the Respiratory Virus Transmission Network-Sentinel, Respiratory Virus Transmission Network-National, and FluTES-C study teams. At Marshfield Clinic Research Institute: Madalyn Palmquist, Lynn Ivacic, Sarah Kopitzke, Erik Kronholm, and Carla Rottscheitt. At Westat: Steph Wraith PhD, Sarah Ezzi BS, and Kathleen Paul BBA. At Columbia: Anny L. Diaz Perez MD, Ana M. Valdez de Romero JD, Michelle Rodriguez BA, and Liqun Wang MS. At the University of North Carolina: Quenla Haehnel, Julienne Reynolds, Amy Yang, Katie Murray, Brittany Figueroa, Anna McShea, Melody Liu, and Miriana Moreno Zivanovich. At the University of Arizona: Mokenge Ndiva Mongoh and Saskia I Smidt. At Stanford: Rosita Thiessen BA CCRP, Alondra A. Aguilar BA, Emma Stainton BS, Grave K-Y. Tam BS, Jonathan Altamirano MS, Leanne X. Chun BBiomed, Rasika Behl MPH, Samantha A. Ferguson BA, Yuan J. Carrington BA, BBA, Frank S. Zhou BS. At Vanderbilt University Medical Center: John Meghreblian, Lauren Milner, Brittany Creasman, Andrea Stafford-Hintz, Ryan Dalforno, Catalina Padilla-Azain, Jorge Celedonio, Afan Swan, Onika Abrams, Judy King, Erica Anderson, Samuel Massion, Daniel Chandler. Paige Yates, Abigail Harris, Brianna Schibley-Laird, Mason Speirs, Sarah Clarke, Alexis Ozden, Marcia Blair, Rendie McHenry, and Erica Anderson.

## Contributions

AMM, HKT, HQM, KGM, EAB, HSI, MR, CR, JED, CO, and CGG contributed to development of the protocol. HKT, HQM, KGM, NMB, KDE, SR, EAB, YM, KL, EA, SD, DO, and CGG were site principal investigators. AL, KEH, EEB, JG, VO, LSM, CYV, AB, JL, LC, GT, ML, KIL, JC, NH, and JS were responsible for data collection, including consent, enrollments, and/or laboratory procedures. AMM, JD, SJ, SSJ, PS, JG, VO, CYV, AB, LC, GT, ML, KIL, KH, YZ, and CL contributed to data management and curation. AMM performed formal analyses and wrote the first draft of the paper. AL, HKT, HQM, SSJ, PS, KGM, NMB, KDE, SR, PS, EAB, YM, KL, EA, MR, and CGG reviewed results and provided substantial critical feedback on interim analyses. All authors read, provided feedback on, and approved the final version of the manuscript, and had final responsibility for the decision to submit for publication. While multiple authors had access to each data source contributing to the study, AMM, JD, SJ, and MR had direct access to all data sources and have independently accessed and verified all underlying data.

## Supplement

**Supplementary Table 1.** Mappings of SARS-CoV-2 variants to PANGO and Nextclade lineages to assigned variant categories among infected household members in a multisite case-ascertained household transmission study, United States, September 2021-September 2022.

| PANGO | Nextclade | Assigned variant | Cases detected<br>among all enrolled<br>participants |
| --- | --- | --- | --- |
| AY.1 | 21J (Delta) | Delta | 1 |
| AY.100 | 21J (Delta) | Delta | 1 |
| AY.103 | 21J (Delta) | Delta | 10 |
| AY.25 | 21J (Delta) | Delta | 2 |
| AY.3 | 21J (Delta) | Delta | 4 |
| AY.44 | 21J (Delta) | Delta | 1 |
| BA.1 | 21K (Omicron) | Omicron BA.1 | 2 |
| BA.1.1 | 21K (Omicron) | Omicron BA.1 | 82 |
| BA.1.1.16 | 21K (Omicron) | Omicron BA.1 | 1 |
| BA.1.1.18 | 21K (Omicron) | Omicron BA.1 | 24 |
| BA.1.15 | 21K (Omicron) | Omicron BA.1 | 5 |
| BA.1.15.2 | 21K (Omicron) | Omicron BA.1 | 1 |
| BA.1.17.2 | 21K (Omicron) | Omicron BA.1 | 10 |
| BA.1.20 | 21K (Omicron) | Omicron BA.1 | 3 |
| BA.2 | 21L (Omicron) | Omicron BA.2 | 46 |
| BA.2.1 | 21L (Omicron) | Omicron BA.2 | 2 |
| BA.2.10.1 | 21L (Omicron) | Omicron BA.2 | 2 |
| BA.2.12 | 21L (Omicron) | Omicron BA.2 | 3 |
| BA.2.12.1 | 21L (Omicron) | Omicron BA.2 | 22 |
| BA.2.12.1 | 22C (Omicron) | Omicron BA.2 | 95 |

| **PANGO** | **Nextclade** | **Assigned variant** | **Cases detected**<br>**among all enrolled**<br>**participants** |
| --- | --- | --- | --- |
| BA.2.13 | 21L (Omicron) | Omicron BA.2 | 3 |
| BA.2.18 | 21L (Omicron) | Omicron BA.2 | 2 |
| BA.2.23 | 21L (Omicron) | Omicron BA.2 | 2 |
| BA.2.3 | 21L (Omicron) | Omicron BA.2 | 15 |
| BA.2.40.1 | 21L (Omicron) | Omicron BA.2 | 2 |
| BA.2.48 | 21L (Omicron) | Omicron BA.2 | 3 |
| BA.2.65 | 21L (Omicron) | Omicron BA.2 | 2 |
| BA.2.68 | 21L (Omicron) | Omicron BA.2 | 1 |
| BA.2.9 | 21L (Omicron) | Omicron BA.2 | 5 |
| BG.2 | 22C (Omicron) | Omicron BA.2 | 1 |
| BA.4 | 21L (Omicron) | Omicron BA.4 | 7 |
| BA.4 | 22A (Omicron) | Omicron BA.4 | 7 |
| BA.4.1 | 21L (Omicron) | Omicron BA.4 | 3 |
| BA.4.1 | 22A (Omicron) | Omicron BA.4 | 15 |
| BA.4.2 | 22A (Omicron) | Omicron BA.4 | 3 |
| BA.4.4 | 21L (Omicron) | Omicron BA.4 | 1 |
| BA.4.6 | 21L (Omicron) | Omicron BA.4 | 3 |
| BA.4.6 | 22A (Omicron) | Omicron BA.4 | 14 |
| BA.5 | 21L (Omicron) | Omicron BA.5 | 1 |
| BA.5 | 22B (Omicron) | Omicron BA.5 | 2 |
| BA.5.1 | 21L (Omicron) | Omicron BA.5 | 12 |
| BA.5.1 | 22B (Omicron) | Omicron BA.5 | 25 |
| BA.5.1.1 | 21L (Omicron) | Omicron BA.5 | 1 |
| BA.5.1.1 | 22B (Omicron) | Omicron BA.5 | 5 |
| BA.5.1.10 | 22B (Omicron) | Omicron BA.5 | 1 |
| BA.5.1.21 | 22B (Omicron) | Omicron BA.5 | 2 |
| BA.5.1.3 | 22B (Omicron) | Omicron BA.5 | 2 |
| BA.5.1.5 | 21L (Omicron) | Omicron BA.5 | 1 |
| BA.5.2 | 21L (Omicron) | Omicron BA.5 | 8 |
| BA.5.2 | 22B (Omicron) | Omicron BA.5 | 28 |
| BA.5.2.1 | 21L (Omicron) | Omicron BA.5 | 19 |
| BA.5.2.1 | 22B (Omicron) | Omicron BA.5 | 65 |
| BA.5.2.20 | 22B (Omicron) | Omicron BA.5 | 2 |
| BA.5.2.9 | 22B (Omicron) | Omicron BA.5 | 3 |
| BA.5.3.1 | 22B (Omicron) | Omicron BA.5 | 2 |
| BA.5.5 | 21L (Omicron) | Omicron BA.5 | 11 |
| BA.5.5 | 22B (Omicron) | Omicron BA.5 | 45 |
| BA.5.6 | 21L (Omicron) | Omicron BA.5 | 4 |
| BA.5.6 | 22B (Omicron) | Omicron BA.5 | 27 |
| BA.5.6.2 | 22B (Omicron) | Omicron BA.5 | 2 |
| BA.5.8 | 22B (Omicron) | Omicron BA.5 | 2 |
| BE.1 | 22B (Omicron) | Omicron BA.5 | 3 |
| BE.1.1 | 22B (Omicron) | Omicron BA.5 | 6 |
| BE.2 | 22B (Omicron) | Omicron BA.5 | 1 |
| BE.3 | 22B (Omicron) | Omicron BA.5 | 14 |
| BF.10 | 22B (Omicron) | Omicron BA.5 | 8 |
| BF.13 | 21L (Omicron) | Omicron BA.5 | 1 |
| BF.13 | 22B (Omicron) | Omicron BA.5 | 2 |
| BF.21 | 22B (Omicron) | Omicron BA.5 | 2 |
| BF.25 | 22B (Omicron) | Omicron BA.5 | 2 |
| BF.26 | 22B (Omicron) | Omicron BA.5 | 5 |
| BF.5 | 21L (Omicron) | Omicron BA.5 | 1 |
| BF.5 | 22B (Omicron) | Omicron BA.5 | 6 |
| BF.8 | 22B (Omicron) | Omicron BA.5 | 1 |

**Supplementary Figure 1.**
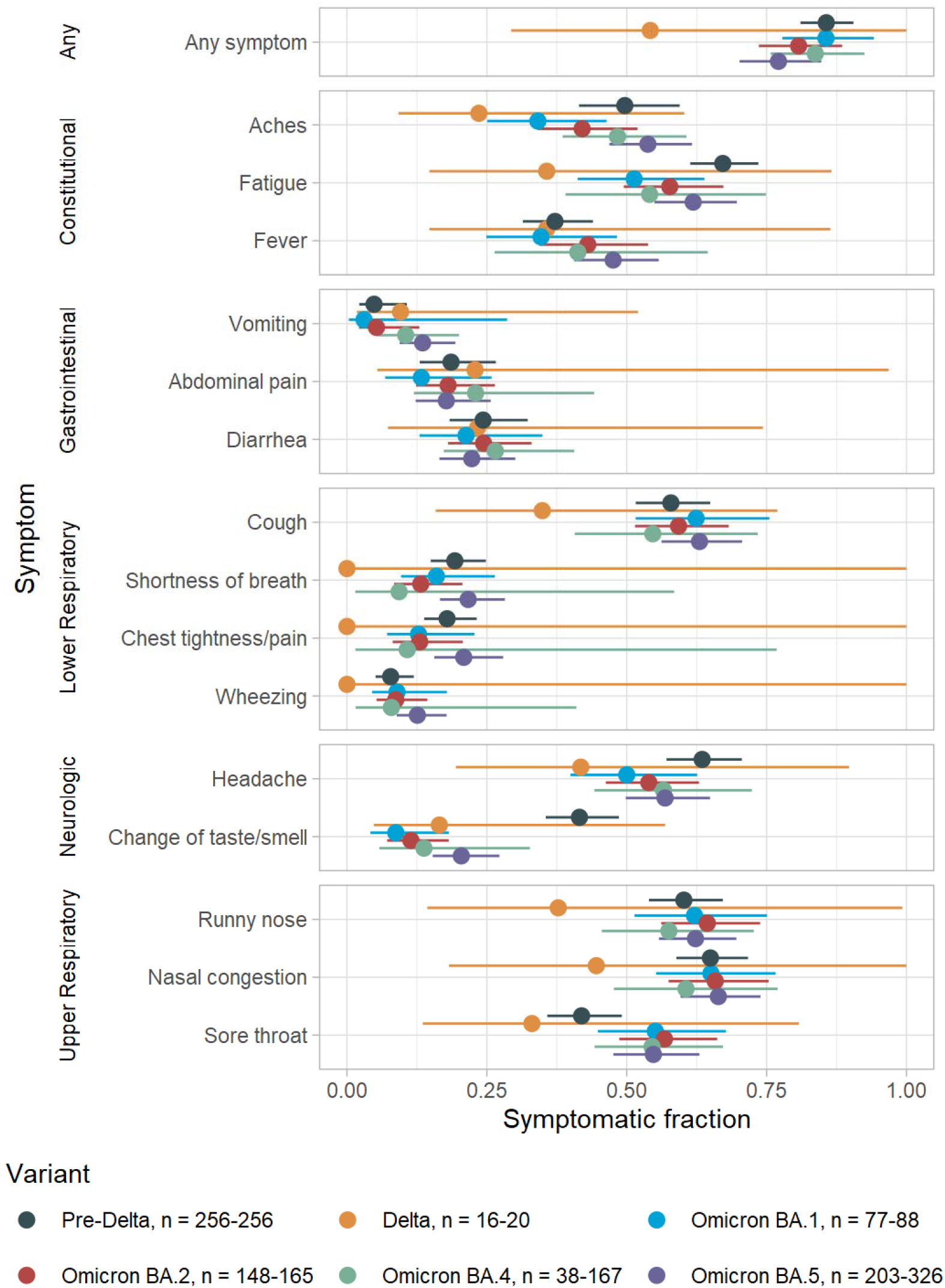
Symptomatic fraction of infected household contacts across six SARS-CoV-2 variants, United States, 2020-2022. Variant indicates the SARS-CoV-2 variant detected in the household for each infected household contact. Sample sizes denote the range of infected household contacts for each variant across five multiply imputed datasets, where imputation was used to address missingness in sequencing data.

**Supplementary Table 2.**
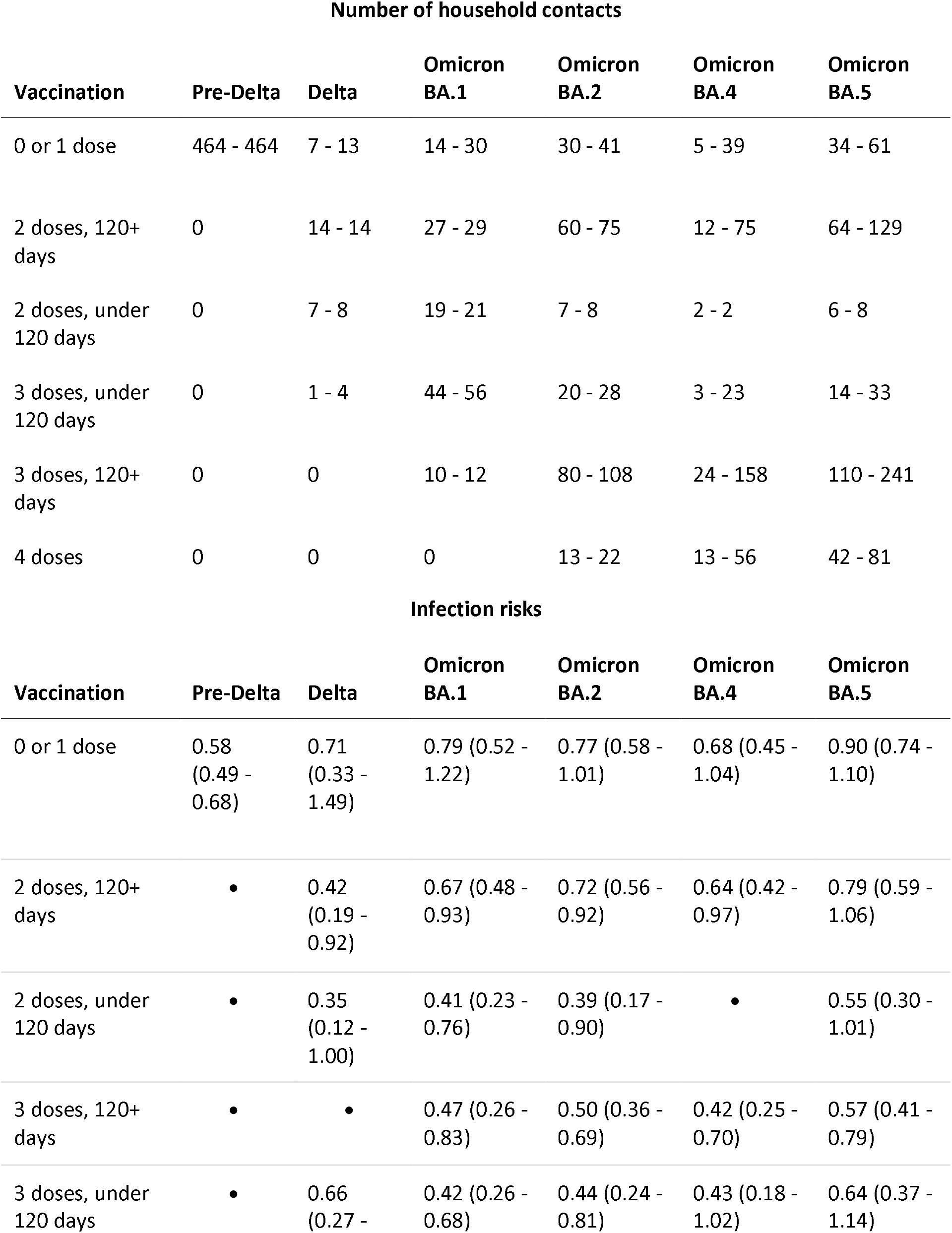
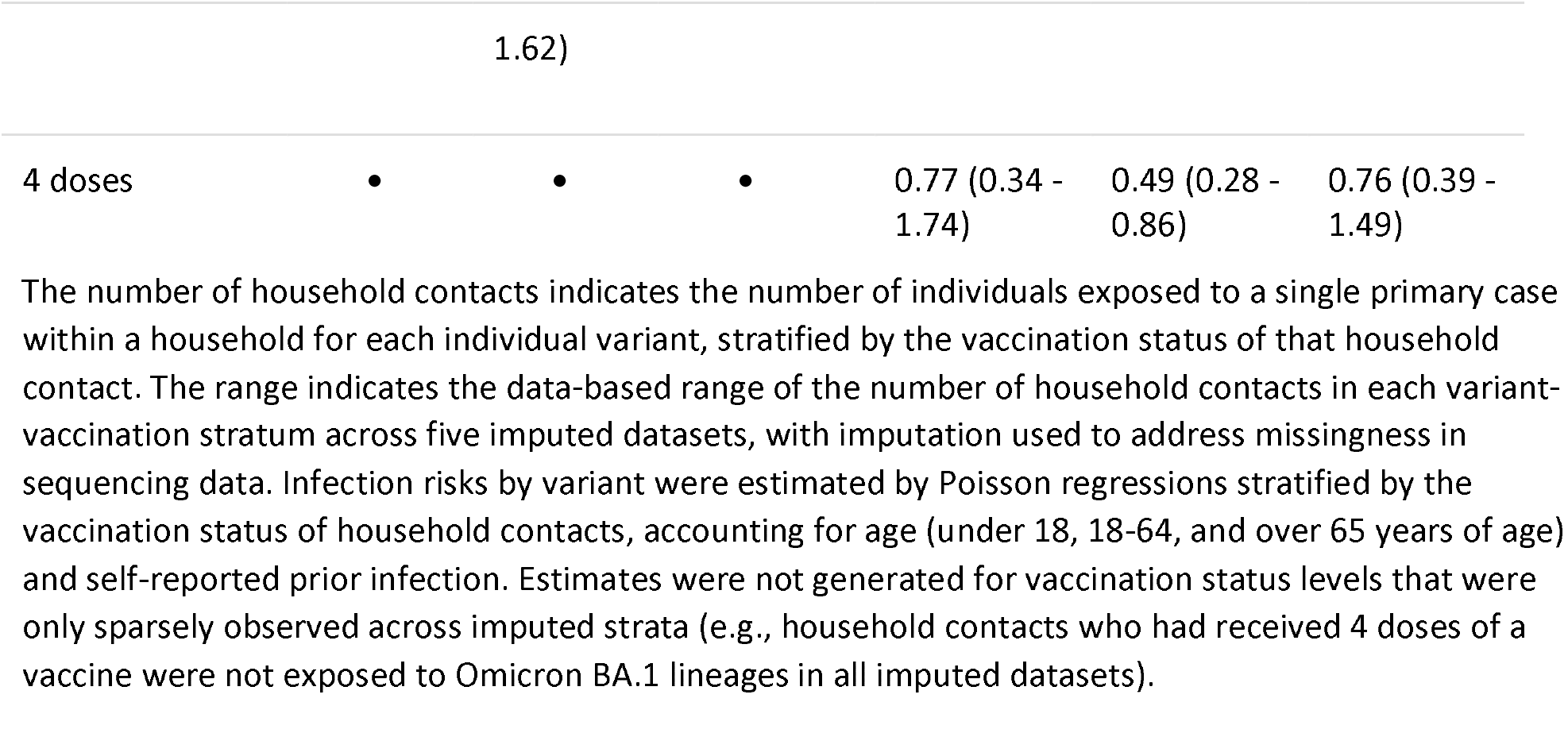
Number of household contacts and infection risks among household contacts, by SARS-CoV-2 variant and vaccination status, United States, 2020-2022.

**Supplementary Figure 2.**
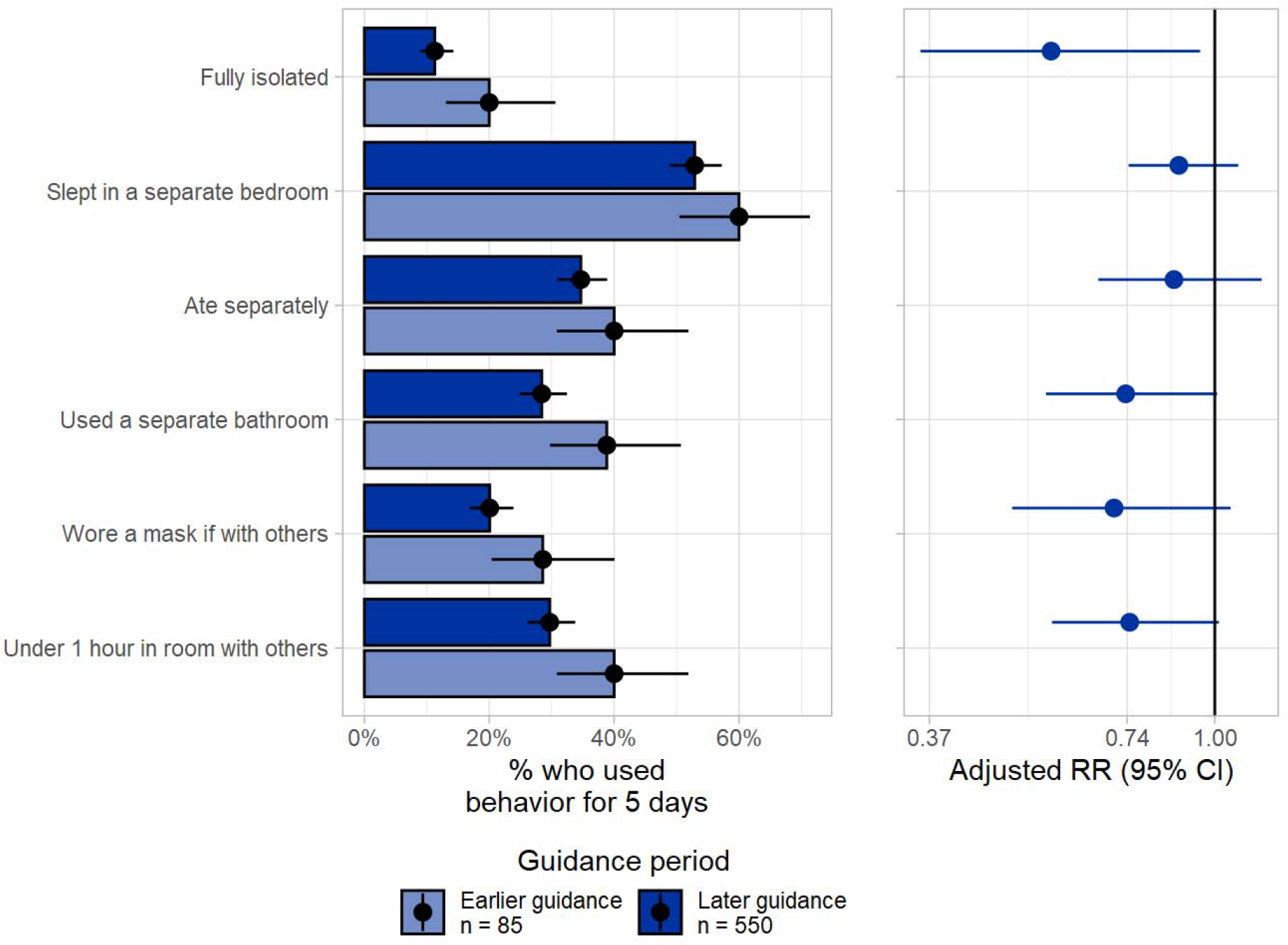
Within-household infection control practices of primary cases across SARS-CoV-2 variant periods, United States, November 2020 – September 2022. The period in which earlier guidance was assessed began in November 2020, and ended December 26, 2021, and the later guidance period began December 27, 2021 and ended in September, 2022. In the earlier guidance period, cases were recommended to isolate from others for ten full days from symptom onset. In the later guidance period, cases were recommended to isolate from others for five days, and to wear a mask as an additional infection control practice for an additional five days. Here, fully isolated indicates that the individual slept in a separate bedroom, ate separately, used a separate bathroom, spent under an hour in the same room as others, and wore a mask around others if spending time together every day for five days, starting from the day after symptom onset. For relative ratios, the reference group was primary cases with symptom onset beginning in the earlier guidance period.

